# Pulmonary Hypertension Classification using Artificial Intelligence and Chest X-Ray:ATA AI STUDY-1

**DOI:** 10.1101/2023.04.14.23288561

**Authors:** Tarık Kıvrak, Burcu Yagmur, Hilal Erken, Derya Kocakaya, Turker Tuncer, Şengül Doğan, Orhan Yaman, Umit Yasar Sinan, Sena Sert Sekerci, Cagri Yayla, Ufuk Iyigun, Mehmet Kis, Ozkan Karaca, Emrah Yesil, Elif Ilkay Yuce Ersoy, Bahar Tekin Tak, Ahmet Oz, Mehmet Kaplan, Zeynep Ulutas, Gamze Yeter Aslan, Nihan Kahya Eren, Fatma Nihan Turhan Caglar, Hatice Solmaz, Ozge Ozden, Hakan Gunes, Umut Kocabas, Mustafa Yenercag, Omer Isık, Cem Yesilkaya, Ali Nail Kaya, Sefa Erdi Omur, Anil Sahin, Erdal In, Nurcan Kırıcı Berber, Cigdem Ileri Dogan, Fatih Poyraz, Emin Erdem Kaya, Ayca Gumusdag, Omer Kumet, Hakki Kaya, Remzi Sarikaya, Seda Turkan Tan, Hidayet Ozan Arabaci, Rengin Cetin Guvenc, Mehtap Yeni, Burcak Kılıckıran Avci, Dilek Cicek Yilmaz, Ahmet Celik, Berkay Ekici, Aycan Fahri Erkan, Veysel Ozgur Baris, Taner Seker, Ferit Böyük, Mehmet Mustafa Can, Hasan Gungor, Hakki Simsek, Bedrettin Yildizeli, Mehmet Ali Kobat, Mehmet Akbulut, Mehdi Zoghi, Omer Kozan

**Affiliations:** Department of Cardiology, Firat University Hospital, Firat University, Elazig, Turkey; Department of Digital Forensics Engineering, College of Technology, Firat University, Elazig, Turkey; Department of Cardiology, Ankara Etlik State Hospital, Ankara, Turkey; Department of Chest Disease, Marmara University Hospital, Marmara University, Istanbul, Turkey; Department of Cardiology, Cardiology Institute, İstanbul, Turkey; Department of Cardiology, Siyami Ersek Hospital, Istanbul, Turkey; Department of Cardiology, Ankara State Hospital, Ankara, Turkey; Department of Cardiology, Hatay State Hospital, Hatay, Turkey; Department of Cardiology, Dokuz Eylul University Hospital, Dokuz Eylul University, Izmir, Turkey; Department of Cardiology, Gaziantep State Hospital, Gaziantep, Turkey; Department of Cardiology, Mersin University Hospital, Mersin University, Mersin, Turkey; Department of Cardiology, Istanbul Education and Research Hospital, Istanbul, Turkey; Department of Cardiology, Gaziantep University Hospital, Gaziantep University, Gaziantep, Turkey; Department of Cardiology, Inonu University Hospital, Inonu University, Malatya, Turkey; Department of Cardiology, Kepez State Hospital, Firat University, Antalya, Turkey; Department of Cardiology, Izmir Katip Celebi University Hospital, Katip Celebi University, Izmir, Turkey; Department of Cardiology, Bakirkoy Education and Research Hospital, Istanbul, Turkey; Department of Cardiology, Tepecik Education and Research Hospital, Izmir, Turkey; Department of Cardiology, Bahcelievler Memorial Hospital, Istanbul, Turkey; Department of Cardiology, Sutcu Imam University Hospital, Sutcu Imam University, Maras, Turkey; Department of Cardiology, Baskent University Hospital,Baskent University, Izmir, Turkey; Department of Cardiology, Ordu University Hospital, Ordu University, Ordu, Turkey; Department of Cardiology, Adnan Menderes University Hospital, Adnan Menderes University, Aydın, Turkey; Department of Cardiology, Ahi Evran University Hospital, Ahi Evran University, Trabzon, Turkey; Department of Cardiology, Gaziosmanpasa University Hospital, Gaziosmanpasa University, Tokat,Turkey; Department of Cardiology, Cumhuriyet University Hospital, Cumhuriyet University, Sivas, Turkey; Department of Cardiology, Turgut Ozal University Hospital, Turgut Ozal University, Malatya, Turkey; Department of Cardiology, Kosuyolu Research and Education Hospital, Istanbul; Department of Cardiology, Bagcılar Education and Research Hospital, Istanbul, Turkey; Department of Cardiology, Van Education and Research Hospital, Van, Turkey; Department of Cardiology, Ankara University Hospital, Ankara University, Ankara, Turkey; Department of Cardiology, Okan University Hospital, Okan University, Istanbul, Turkey; Department of Cardiology, Isparta Education and Research Hospital, Isparta, Turkey; Department of Cardiology, Cerrahpasa Medical School Hospital, Istanbul University, Istanbul, Turkey; Department of Cardiology, Ufuk University Hospital, Ufuk University, Istanbul, Turkey; Department of Cardiology, Adana Education and Research Hospital, Adana, Turkey; Department of Cardiology, Yedikule Thoracic Disease and Thoracic Surgery Training and Research Hospital, Istanbul, Turkey; Department of Cardiology, Dicle University Hospital, Dicle University, Diyarbakir, Turkey; Department of Thoracic Surgery, Marmara University Hospital,Marmara University, Istanbul, Turkey; Department of Cardiology, Baskent University Hospital, Baskent University, Istanbul, Turkey

**Keywords:** pulmonary hypertension classification, artificial intelligence, Chest X-Ray

## Abstract

An accurate diagnosis of pulmonary hypertension (PH) is crucial to ensure that patients receive timely treatment. One of the used imaging models to detect pulmonary hypertension is the X-ray. Therefore, a new automated PH-type classification model has been presented to depict the separation ability of deep learning for PH types. We retrospectively enrolled 6642 images of patients with PH and the control group. A new X-ray image dataset was collected from a multicentre in this work. A transfer learning-based image classification model has been presented in classifying PH types. Our proposed model was applied to the collected dataset, and this dataset contains six categories (five PH and a non-PH). The presented deep feature engineering (computer vision) model attained 86.14% accuracy on this dataset. According to the extracted ROC curve, the average area under the curve rate has been calculated at 0.945. Therefore, we believe that our proposed model can easily separate PH and non-PH X-ray images.

## 1. Introduction

An accurate diagnosis of pulmonary hypertension (PH) is crucial to ensure that patients receive timely treatment for a progressive clinical course. [1] Although approaches for precise diagnosis of PH may avoid the development of symptomatic heart failure, a low-cost and non-invasive screening tool does not exist in the clinical setting. As a result, several groups have sought to identify minimally invasive or non-invasive approaches to identifying patients with PH. For example, the American College of Chest Physicians has recommended obtaining a chest X-ray (CXR) in patients suspected of having PH to reveal features supportive of PH diagnosis [2]. However, it is well known that the sensitivity and specificity are low. Currently, the recommended test for screening is echocardiography; however, the test requires intensive training and highly qualified technical staff and is relatively expensive [3]. In addition, although the multi-modality assessment plays a central role in evaluating PH [4], the accessibility of diagnostic modalities is sometimes limited in many remote areas and facilities.

Artificial intelligence (AI) has been applied to recognize subtle patterns in digital data in medical fields [5,6]. For example, a family of algorithms has led to state-of-the-art improvements in word recognition, visual object recognition, object detection, etc. [7,8]. Recently, simple digital data (e.g., electrocardiogram) can identify asymptomatic left ventricular dysfunction at baseline and during follow-up using AI [9]. Thus, we hypothesized that the application of AI to the CXR could identify a patient with PH, and it is possible to determine which group it is in. We created, trained, validated, and then tried AI models to test this hypothesis.

## 2. Materials and Methods

### Study population

We retrospectively enrolled a total of 2005 patients with chest CXR referred to our laboratory to evaluate pulmonary hypertension classification. These consecutive patients had experienced symptoms or signs of pulmonary hypertension in our clinics. In our study, the findings for PH were observed in CXR (1: frontal chest radiograph shows a prominent central pulmonary artery, 2: dilated right interlobar artery, and 3: pruning of peripheral pulmonary vascularity). We excluded patients with pacemakers, scoliosis, infiltrative pneumonia, and electrode attached. The collected dataset comprises 6662 X-ray images with six categories. These categories are given as follows: (1) pulmonary arterial hypertension, (2) pulmonary hypertension due to left heart disease, (3) pulmonary hypertension due to lung disease and hypoxia, (4) chronic thromboembolic pulmonary hypertension, (5) pulmonary hypertension with unclear and multi-factorial mechanism and (6) non-PH. The collected images have been stored in JPG format. The attributes of the images are tabulated in Table 1, which organizes the first five classes from different medical centers. What downloaded the sixth class (healthy class) from Kaggle [10] dataset, and professional cardiologists selected non-PH X-rays images from this dataset [11-12]

**Table 1.** Attributes of the X-Ray image dataset collected.

| No | Category | Number of images |
| --- | --- | --- |
| 1 | Pulmonary arterial hypertension | 2563 |
| 2 | Pulmonary hypertension due to left heart disease | 891 |
| 3 | Pulmonary hypertension due to lung disease and hypoxia | 330 |
| 4 | Chronic thromboembolic pulmonary hypertension | 670 |
| 5 | Pulmonary hypertension with unclear and multi-factorial mechanisms | 65 |
| 6 | Non-PH | 2145 |
| Total |  | 6642 |

The Institutional Review Board of the Firat University Hospital approved the study protocol. All patients signed informed consent. All methods were by the relevant guidelines and regulations.

### Eyeball assessment

Visual assessments were interpreted by the consensus of 4 physicians who were blinded to medical history data using Chest X-ray images based on the radiographic findings of PH: (1) frontal chest radiograph shows a prominent central pulmonary artery, (2) dilated right interlobar artery and (3) pruning of peripheral pulmonary vascularity in the guideline and report [11].

### Import data

Each case contains Chest X-Ray images. We transformed all DICOM images into 512 × 512 resolution portable network graphic images with down-sampling. All data were divided into six groups and were used as training and validation to create a model.

### Deep learning model

The main objective of the proposed model is to extract the most informative features from an X-Ray image with a low time burden. Moreover, patch-based in-depth feature extraction methods/deep models, for instance, MLP-mixer [13] or vision transformers (ViT)[14], have high image classification ability. However, these models generate many patches, which increases the time complexity of the models. This work uses a new patch division model to decrease the times’ complexity since this model generates fewer patches to extract features. The pretrained EfficientNetb0 [15] has been utilized as a deep feature generator. EfficientNetb0 was trained using the ImageNet 1K datasets. This dataset contains approximately 1.2 million images with 1000 classes. Two layers of this network (EfficientNetb0) have been used to extract deep features. The features created from each patch have been merged. Therefore, this model is called NPEffNetb0. INCA has been applied to choose the best feature vector automatically. In the classification phase. SVM classifier with a 70:30 split ratio has been used. The visual denotation of the NPEffNetb0 feature extraction-based PH classification model is depicted in Figure 1.

**Figure 1.**
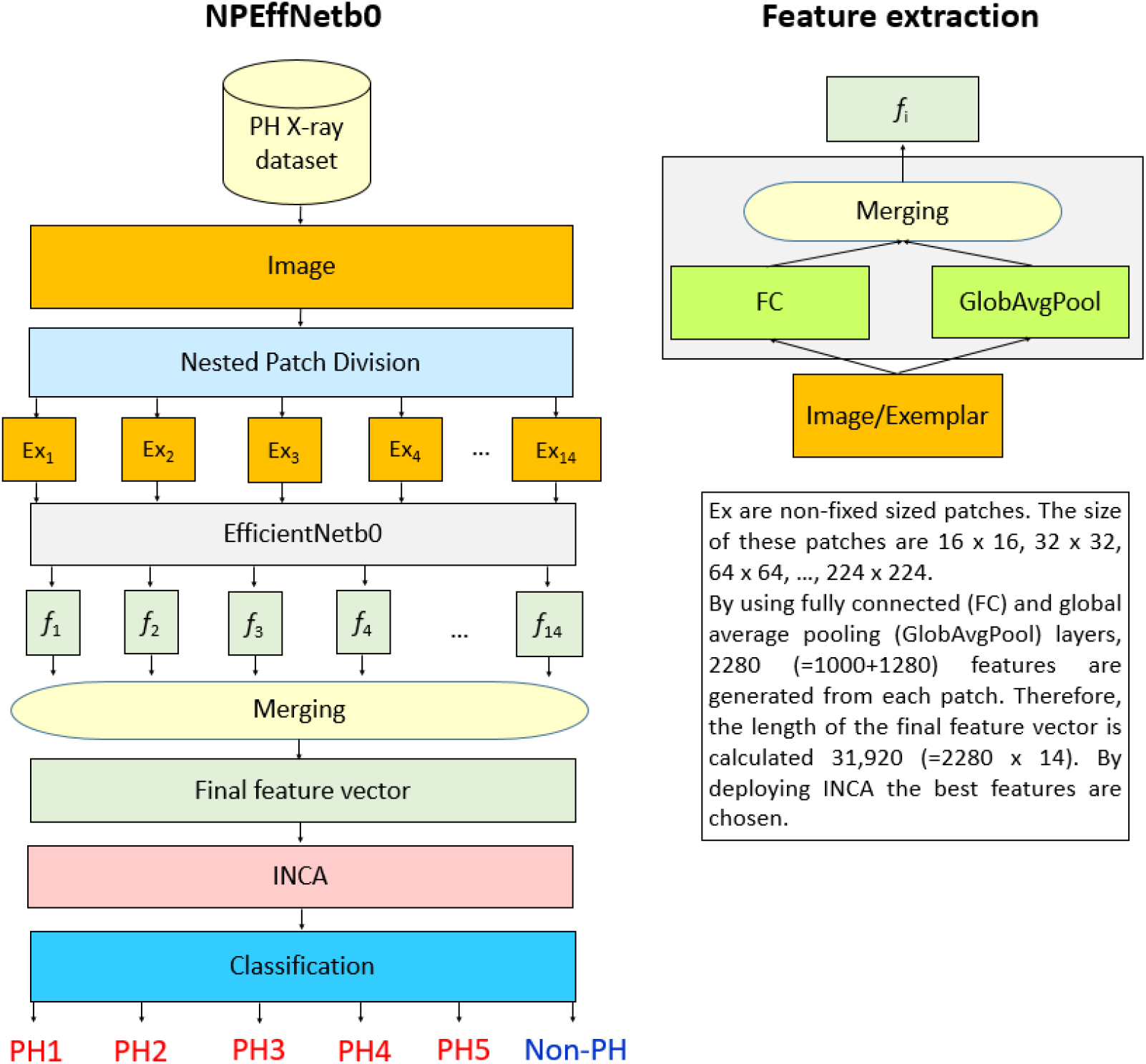
Graph of the presented NPEffNetb0 classification model.

The steps of the presented NPEffNetb0 feature extraction-based PH classification model are:

Step 1: Resize each image into 224 × 224 sized images.

Step 2: Deploy nested patch (NP) division model to extract non-fixed 14 patches. The algorithm of the NP division is given in Algorithm 1.

Step 3: Extract features from each patch using fully connected and global average pooling layers of the pretrained EfficientNetb0.

Step 4: Merge the generated 14 feature vectors to obtain the final vector.

Step 5: Apply INCA [16] to choose the best feature vector. Herein, SVM has been considered an error/misclassification rate. Therefore, in this research, the length of the optimal feature vector is calculated as 829.

Step 6: Classify the chosen 829 features by deploying an SVM [17-18] classifier. The used SVM classifier is called Cubic SVM. The properties of the used Cubic SVM [19] are:

Kernel function: Polynomial, Polynomial

order: Three, Kernel scale: Automatic,

Box constraint level: one,

Validation: Held-out validation with a split ratio of 70:30.

Using X-ray images, these steps define the proposed NPEffNetb0 feature extraction-based PH classification model.

## 3. Results

### 3.1. Performance evaluation

We have used some parameters. We have chosen the most used four performance evaluation metrics in this work. These are accuracy (the most preferred performance metric and it shows the prediction performance of the used model), recall (to get class-wise/ailment-wise accuracy), precision (it is a critical parameter for biomedical datasets since the precision rate of the accurate predicted/diagnosed and falsely expected/diagnosed) and F1-score (harmonic mean/average of the precision and recall). Therefore, accuracy, recall, precision, and F1-score were considered to evaluate the performance of the proposed approach. The explanations of these metrics are mathematically given in Eqs. 1-4.

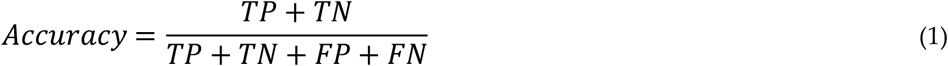

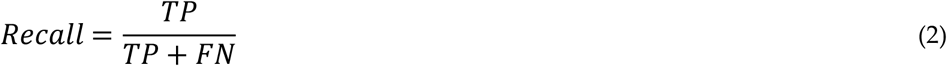

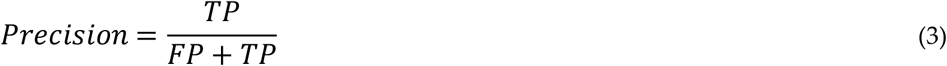

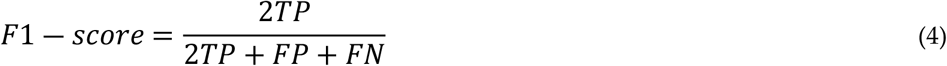

where TP, TN, FP, and FN are true positives, true negatives, false positives, and false negatives.

### 3.2. Experimental Results

This work applied a new classification model to the collected PH X-ray image dataset. The used dataset was collected from different medical centers in JPG format. Who used MATLAB programming tool to develop the proposed NPEffNetb0. This work implemented a supervised learning application using the used dataset and the presented NPEffNetb0.

The used dataset contains six classes. Hence, both overall and class-wise results have been calculated in this research. We generated a confusion matrix to calculate these results, and this confusion matrix is depicted in Figure 2.

**Figure 2.**
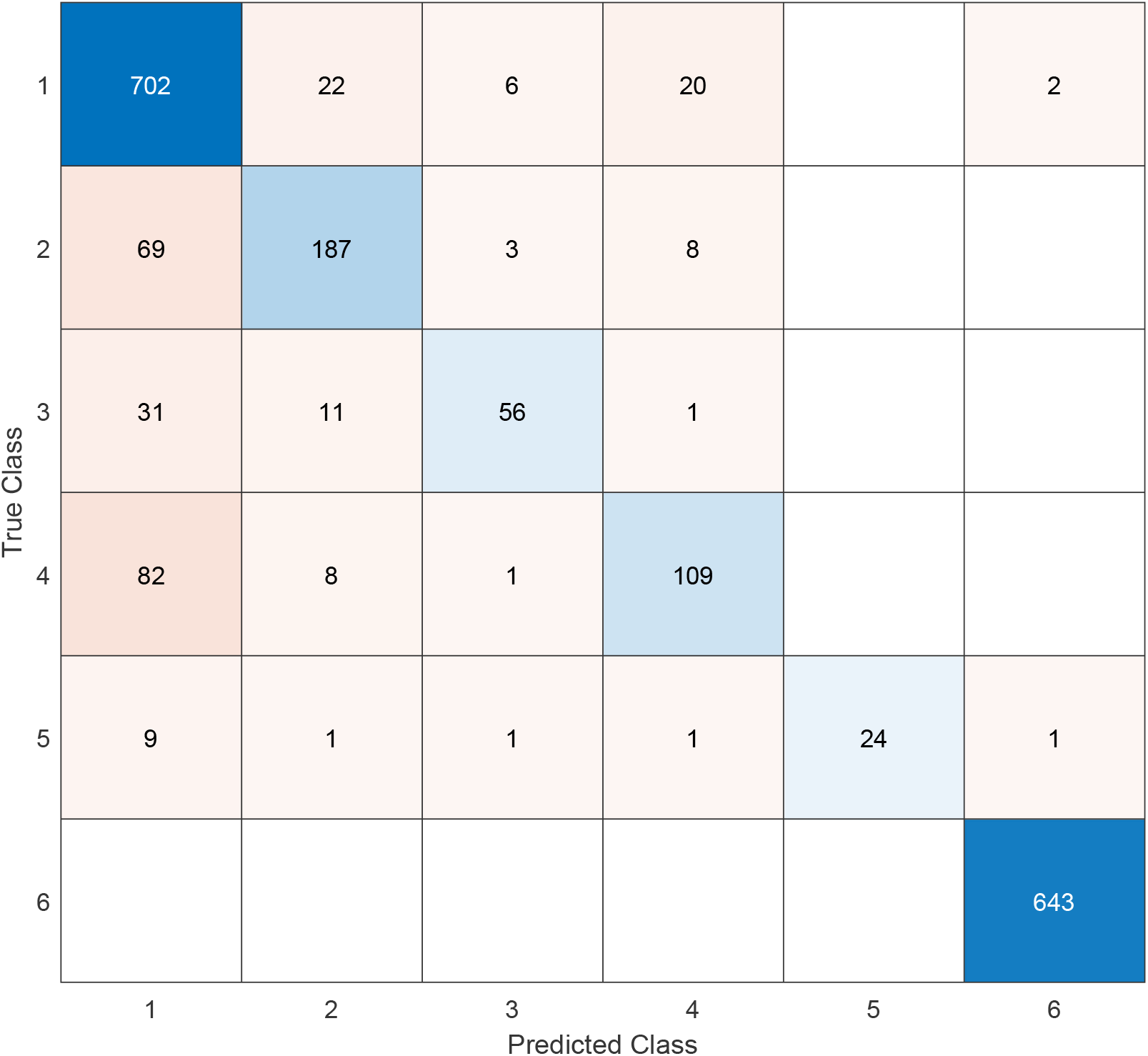
Confusion matrix calculated of the proposed EffNetb0 on the collected X-ray dataset for PH detection.

By using this confusion matrix (see Figure 2), average precision (AP), unweighted average recall (UAR), and overall F1-score (F1) performances [20-22] have been calculated, and the calculated overall results have been tabulated in Table 2. Class-wise results of the proposed model have been calculated, and the results are denoted in Figure 3. Figure 3 demonstrated that the presented NPEffNetb0 feature extraction-based model attained a 100% recall value for the sixth class. It is suggested that our model successfully separates X-ray with PH and X-ray with non-PH. The worst recall is attained in the fourth class, and the recall value of this class is equal to 54.50%. Moreover, the Receiver Operating Characteristic (ROC) curve of the calculated result has been demonstrated in Figure 4. According to the extracted ROC curve, the average area under the curve rate has been calculated at 0.945.

**Table 2.**
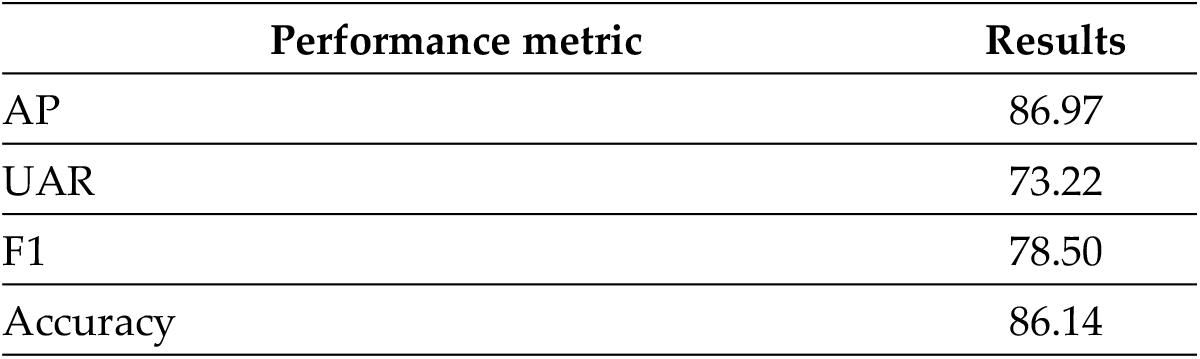
Overall classification performances.

**Figure 3.**
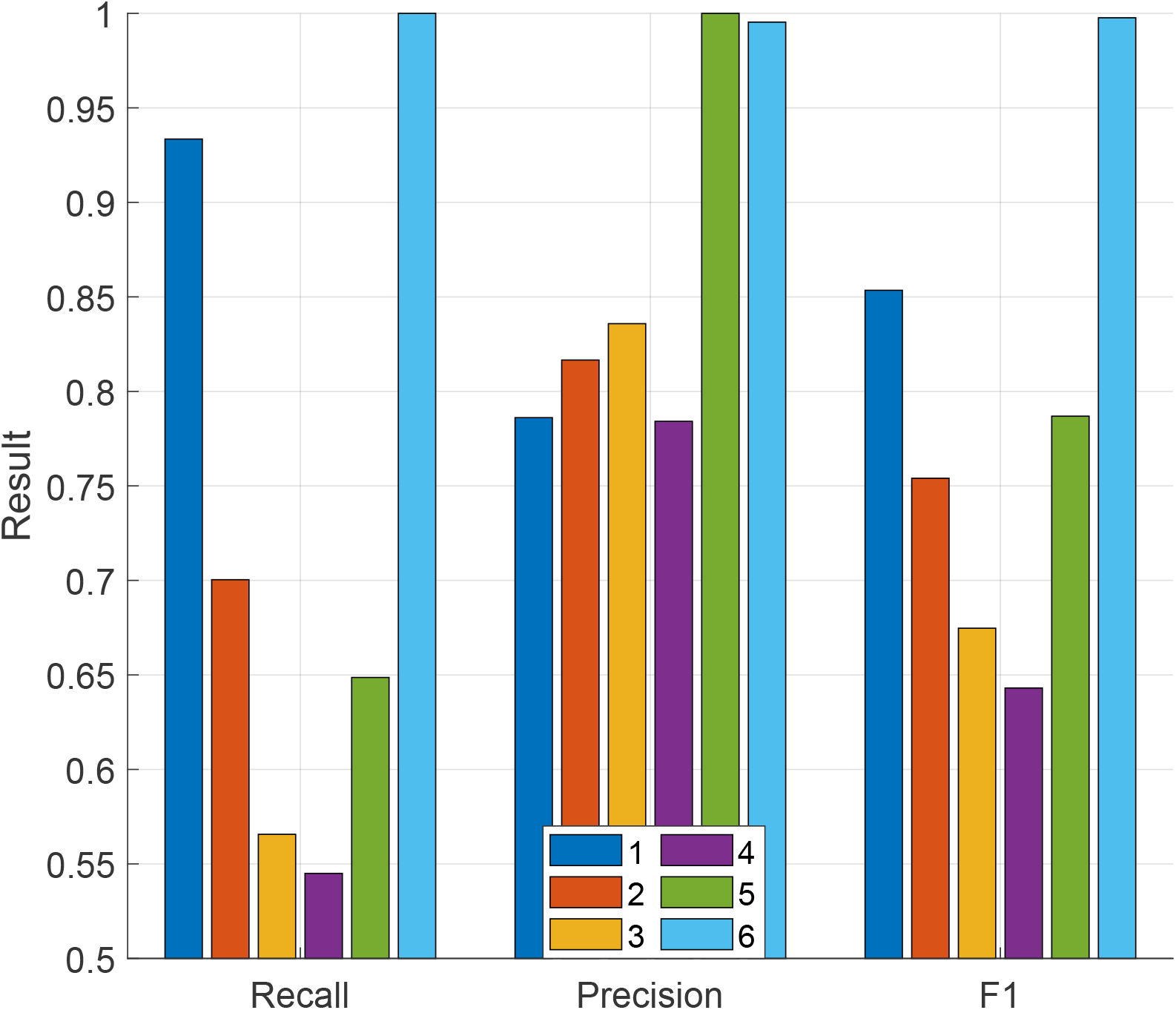
Class-wise performance metrics of the proposed EffNetb0 feature extraction-based model

**Figure 4.**
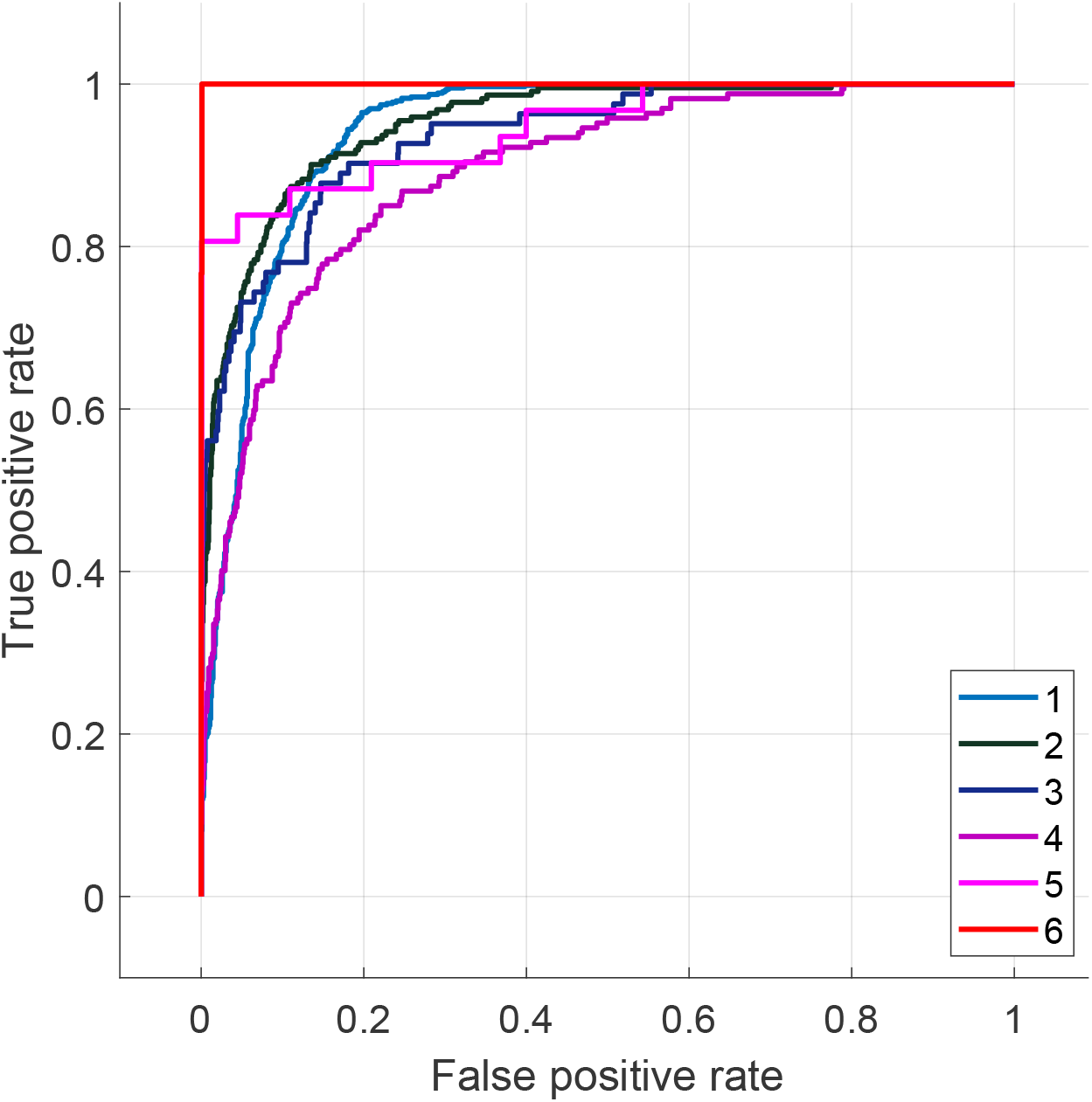
ROC curve of the presented model.

## 4. Discussion

Previously, Chest X-Ray was a simple and economical method available globally. A recent study showed that measurements could identify more subjects suffering from undiagnosed PH [23]. However, the AUC was limited to 0.60–0.62. Moreover, several methods included laboratory data, electrocardiograms, and physical examinations to detect PH [24–27]. In reproducibility, automated assessment is needed to obtain quantitative results without any user interaction, including measurements. Our results demonstrate that it can train an AI model to estimate PH on Chest X-Ray images. We believe this study is a pilot study leaning towards the possibility of applying a DL algorithm in the clinical assessment of pulmonary hypertension. PH type classification is a necessary process for cardiology, and one of the imaging methods for PH is X-ray imaging. This work tested the PH-type classification ability of a deep feature engineering model on an unbalanced X-ray image dataset. The NP model has been used to use the positive effects of patch-based feature extraction. Pretrained EfficientNetb0 was used. To select this function, we evaluated the performances of the eight pre-trained networks. These eight networks (ResNet18 [23] ResNet50 [23], ResNet101 [23], DarkNet19 [24], MobileNetV2 [25], DarkNet53 [24], Xception [26], and EfficientNetb0 [15] were utilized as feature extraction function in our proposed architecture and the calculated classification accuracies have been denoted in Figure 5.

**Figure 5.**
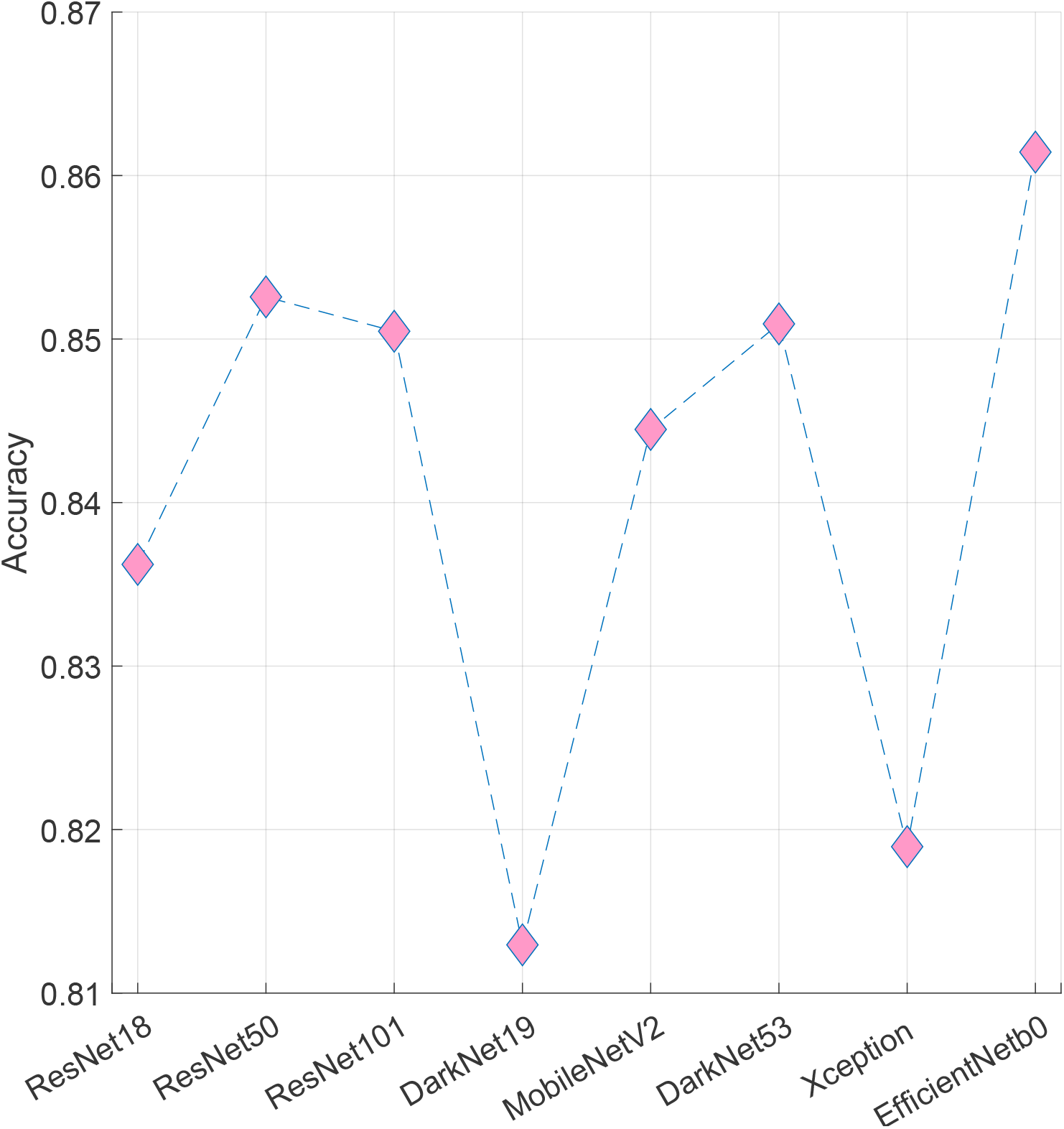
Classification performances (accuracy) of eight pretrained CNNs.

As shown in Figure 5, the best-trained CNN [27] is EfficinetNetb0 since it attained 86.14% classification accuracy. The second CNN is ResNet50 which yielded 85.29% classification accuracy. Therefore, EfficientNetb0 is selected as this architecture’s primary feature extraction function.

The salient/essential points of the proposed model are.

- A novel deep feature engineering model has been presented, and this model attained 86.14% classification accuracy on the collected unbalanced X-ray image dataset.
- We are the first group to use Chest X-Ray images to classify PH categories. Therefore, we collected a relatively large dataset to test our model in this aspect.
- By using NP, both positive effects of the patch-based models have been used, and what implemented this process (feature extraction process) with a short execution time?
- Our proposed NPEffNetb0 feature extraction-based model yielded satisfactory performance for the PH type detection. Furthermore, this model attained recall in some classes (12-14) since this dataset is unbalanced and the used PH type is rarely seen.
- Our results suggest that the AI algorithm can obtain new insight using a standard economical method. Furthermore, the first study demonstrates that the AI algorithm adds further information to Chest X-Ray for PH classification.
- **Comparison with previous analysis**. Previously, Chest X-Ray was an economical and straightforward method available globally. Thus, this technique is a helpful tool for checking patients with suspected elevated PAP [28]. In addition, a recent study showed measurements of Chest X-Ray could lead to identifying more subjects suffering from undiagnosed PH [29]. However, the AUC was limited to 0.60–0.62.
- Moreover, several methods included laboratory data, electrocardiograms, and physical examinations to detect PH [30-33]. In reproducibility, automated assessment is needed to obtain quantitative results without any user interaction, including sizes. Our results demonstrate that it can train an AI model to estimate PH on Chest X-Ray images. We believe this study is a pilot study leaning towards the possibility of applying a DL algorithm in the classification of pulmonary hypertension.
- **Additional knowledge from artificial intelligence for Chest X-rays**. The specific Chest X-Ray characteristics used by the convolutional neural network to classify individuals as having PH are not well known because of a “black box” algorithm. We suspect it is detecting the known Classification of Pulmonary Arterial Hypertension on the Chest X-Ray. According to the results of heat map analysis, AI assessment mainly focused on the heart and lung areas. The AI algorithm might be tracing the process of human knowledge. It may extend the use of AI beyond the capacity of human knowledge in the future.
- **Clinical implications**. Assessment of PH using the AI algorithm is an objective method, and its discrepancy was similar to that of assessment by experts. Chest X-Ray can be used as an inexpensive, standardized, universal test. If many patients can access an inexpensive, reasonable test for PH, individual patients could benefit from early effective therapies. This method may allow us to identify more patients with possible PH in regions lacking sufficient imaging facilities. It is challenging to detect post-capillary PH caused by left heart failure in the clinical setting. On the other hand, early detection of pre-capillary PH is also essential because it is often misdiagnosed, and treatment of pre-capillary PH may differ from post-capillary PH. Future studies involving more significant numbers are required to assess the AI models for the differentiation of pre-and post-capillary PH.
- This study, İt tested the AI tool in patients who have already had an echocardiographic study and Chest X-Ray indicating suspected PH. Because it is impossible to use the right heart catheterization for patients without suspected cardiovascular diseases due to ethical issues, thus, we could not claim any value of the AI assessment in screening patients for PH based on clinical signs alone. However, AI assessment may be considered an option to check the necessity.

## 5. Conclusions

Applying AI to the Chest X-Ray (a conventional, universal, low-cost test) is a potential tool to detect PH. A new deep learning model has been presented to classify PH types in this research. A deep transfer learning-based feature engineering model has been proposed in this research. A new non-fixed size patch-based deep feature extractor has been proposed in this work. The pretrained EfficientNetb0 has been applied to each patch and extracted deep features. The proposed AI model is a deep feature engineering model. Therefore, it selected the features generated with an iterative feature selector (INCA), and these features were classified using an SVM classifier. This is the first automated PH detection model using six deep-learning classes. Our model reached 86.97% average/overall precision, 73.22% UAR, 78.50% overall F1-score, and 86.14% classification accuracy for six classes. Moreover, our proposed nested patch division-based model can easily separate healthy, and it attained PH classes since 100% class-wise recall in the healthy (6th) class.

Our proposed model can separate PH and non-PH X-ray images easily. However, this preliminary work suggests that applying AI to the Chest X-Ray in PH groups has limited performance in some groups. Therefore, we should consider that it is premature to include this technology in the current guidelines.

## Data Availability

yes

